# Shared genetic aetiology between childhood intelligence and longevity

**DOI:** 10.1101/2021.02.10.21251491

**Authors:** W. David Hill, Ian J. Deary

## Abstract

**Background:** Intelligence and longevity are phenotypically and genetically correlated. Whereas molecular genetic data has been used to show that adult intelligence is genetically correlated with longevity, no such analysis has examined the association between childhood intelligence and longevity.

**Method and Results:** Using genome wide association study data on childhood intelligence (n = 12,441) and on parental longevity (n = 389,166) we found a positive genetic correlation of *r*_*g*_ = 0.35 (SE = 0.14, P = 0.01) between childhood intelligence and parental longevity.

**Conclusion:** These results add to the weight of evidence that the phenotypic link between childhood intelligence and longevity is, partly, accounted for by shared genetic aetiology.

## Introduction

Long-term follow-up data from large cohorts sourced from the UK,^1^ Denmark,^2^ Israel,^3^ and Sweden,^4^ have been used to show that higher scores on intelligence tests in youth (childhood, adolescence or young adulthood) are associated with lower risk of mortality from all causes by mid to late adulthood. A systematic review was conducted using 16 separate studies, drawing data from over 1 million participants (including 22 453 deaths). It identified that, on average, for a 1 standard deviation higher intelligence test score in youth, there was a 24% (95% CI = 23% to 25%) lower risk of death during a follow-up period of between 17 to 69 years^5^. This relationship was not confounded by childhood socioeconomic position, was present across a range of cognitive ability, and was present in both men and women^5^. There was some attenuation by education and the person’s own adult occupational social class.

Molecular genetic data sourced from genome-wide association studies has been used to examine the genetic links between measured intelligence and longevity. This is, in part, due to the availability of large GWAS that were conducted to examine the molecular genetic aetiology of people’s differences in intelligence test scores.^6, 7^ These GWAS data enable a comparison between traits; that is, one may compare the loci that attain genome wide statistical significance in one trait with those that are genome wide significant in other traits, including longevity. Furthermore, a genetic correlation between two traits’ GWASs can be derived to describe the average shared genetic effect between two traits. Such analyses have been conducted on intelligence and measures of longevity; one estimate of the genetic link between intelligence and all-cause mortality is *r*_g_ = 0.37, SE = 0.06, P = 0.009.^6^

However, unlike phenotypic analyses, in which childhood intelligence is used as a predictor of longevity, genetic correlations have, so far, only been derived using intelligence as assessed in adulthood and in older age. Whereas the genetic correlation between intelligence in childhood and intelligence in adulthood/older age is high (*r*_g_ = 0.71, SE = 0.10)^8^, the genetic relationship between intelligence and health outcomes have been noted to be dependent on the age at which intelligence was measured^8^.

The goal of the research field of cognitive epidemiology^9^ is to describe and explain phenotypic associations between intelligence tested in youth (which largely avoids reverse causation) with later-life health and death. Part of the phenotypic childhood intelligence-longevity association might be due to shared genetic factors^10^. However, to date, there has been no estimate for the genetic correlation between childhood intelligence and age at death. In order to fill this lacuna in cognitive epidemiology, we use two GWAS to derive a genetic correlation to examine the genetic relationship between intelligence, as assessed in childhood/young adulthood and longevity.

## Method

Two sets of genome wide association study summary data were used. First, to provide genetic association data for measured intelligence in childhood, data from the Childhood Intelligence Consortium (CHIC)^11^ was used (n = 12,441, age range 6–18). Second, to provide genetic association data for age at death, a GWAS of parental longevity was included (n = 389,166).^12^

A Genetic correlation was derived using bi-variate linkage disequilibrium score (LDSC) regression using the data processing pipeline created by Bulik-Sullivan et al. (2015). LD scores and weights were downloaded from (http://www.broadinstitute.org/~bulik/eur_ldscores/) for use with European populations. SNPs were included if they had a minor allele frequency of > 0.01 and an imputation quality score of > 0.9. Following this, SNPs were retained if they were found in HapMap 3 with MAF > 0.05 in the 1000 Genomes EUR reference sample. Next, indels and structural variants were removed along with strand-ambiguous variants. Additionally, SNPs whose alleles did not match those in the 1000 Genomes were also removed. SNPs within the MHC region on chromosome 6 (26M to 34M) were also removed. The presence of outliers can increase the standard error in LDSC regression; therefore, SNPs where χ^2^ > 80 were also removed.

## Results

Using linkage disequilibrium score regression, the SNP-based heritability of childhood intelligence was 27.3% (SE = 4.7%) and the heritability of longevity was 28.9% (SE = 0.7%). The mean χ^2^ statistic for childhood intelligence was 1.08 and the LDSC intercept was 1.00. For longevity, these metrics were χ^2^ = 1.06 and an LDSC intercept of 1.02. The mean χ^2^ statistic provides information on to the degree to which the GWAS test statistics deviate from that which would be expected under the null hypothesis of no association (i.e. if no SNPs were associated with the trait, childhood intelligence and parental longevity, the mean χ^2^ statistic for each of these data sets would be 1). The LDSC intercept is a metric describing what this inflation in the mean χ^2^ statistic is likely to be due to. Should a GWAS be confounded by population stratification and/or cryptic relatedness, this intercept will exceed 1.The results above indicate that, whereas there is an inflation of the GWAS test statistics for childhood intelligence, 96.9% of this inflation is consistent with polygenicity. For longevity, 72.8% of the inflation was due to polygenicity; however, the intercept was also close to 1, indicating no problematic inflation of the test statistics for reasons other than polygenicity.

With respect to the principal result from the present analysis, the genetic correlation between childhood intelligence and parental age at death was *r*_*g*_ = 0.35 (SE = 0.14, P = 0.01). This indicates that some of the genetic variants associated with higher childhood intelligence are also associated with one’s parents’ living longer.

## Discussion

Here we present evidence that some of the same genetic variants are associated with both measured childhood intelligence and longevity. Whereas previous work has examined the genetic association between longevity and intelligence, samples with respect to intelligence measurement have included mostly mature and older adults^7, 13^. This has hampered efforts to understand the phenotypic links between childhood intelligence and longevity. The current results contribute toward filling this gap in our understanding of cognitive epidemiology, and show that pleiotropy provides a partial explanation for this phenotypic link between intelligence and longevity.

The observation that some shared genetic influence is present between childhood intelligence and longevity is consistent with a number of models of pleiotropy^14^. First, genetic correlations can arise due to horizontal pleiotropy. Horizontal pleiotropy describes instances in which a genetic variant (or group of variants) has two independent effects on the two phenotypes that are genetically correlated. Should horizontal pleiotropy be one of the drivers of this genetic link between intelligence and longevity, then it would be evidence for the “system integrity” hypothesis, i.e. that more intelligent people tend to live longer and in better health due to, in part, to genetic effects producing a body and brain more capable of withstanding environmental insults^15^.

Second, the presence of a genetic correlation can sometimes be explained with vertical pleiotropy. Vertical pleiotropy describes instances where two phenotypes are genetically correlated as a consequence of one phenotype’s being causally related to the second. In this type of pleiotropy, the genetic correlation between childhood intelligence and longevity may have arisen, in part, due to childhood intelligence’s providing entry into environments more likely to be conducive to good health. Specifically, for example, intelligence has been found to be causally related to education^16^ and a higher level of education is associated with a higher level of socioeconomic position and less material privation and a greater tendency toward healthier behaviours and higher health literacy.

Spurious pleiotropy may also explain some of the present results. Spurious pleiotropy occurs where two causal (in this instance one causal for intelligence differences and the second causal for longevity differences) are in linkage disequilibrium with each other resulting in a genetic correlation between childhood intelligence and parental longevity at this locus. However, for two polygenic traits, like intelligence and longevity, it is possible that multiple forms of pleiotropy contribute towards the genetic correlation observed in these data.

The genetic correlation derived herein describes the average shared genetic effect across the autosomes. It does not provide information pertaining to which regions of the genome (beyond the autosomal contributions) contribute to this effect. As such it is of limited utility in understanding the shared biological systems (if any) that are linked to the observed genetic correlation. Additional work is required to identify regions of the genome that drive this genetic correlation^17^.

## Data Availability

All data used are publicly available.

## Acknowledgements

The authors are members of the Lothian Birth Cohorts group at the University of Edinburgh, which is supported by Age UK (Disconnected Mind grant), the Medical Research Council (MR/R024065/1), and the USA’s National Institutes of Health (1RO1AG054628-01A1).

WDH is supported by a Career Development Award from the Medical Research Council (MRC) [MR/T030852/1] for the project titled “From genetic sequence to phenotypic consequence: genetic and environmental links between cognitive ability, socioeconomic position, and health”.

## References

1. Čukić I, Brett CE, Calvin CM, Batty GD, Deary IJ. Childhood IQ and survival to 79: Follow-up of 94% of the Scottish Mental Survey 1947. Intelligence 2017; 63: 45–50.

2. Christensen GT, Mortensen EL, Christensen K, Osler M. Intelligence in young adulthood and cause-specific mortality in the Danish Conscription Database – A cohort study of 728,160 men. Intelligence 2016; 59: 64–71.

3. Twig G, Tirosh A, Derazne E, Haklai Z, Goldberger N, Afek A et al. Cognitive function in adolescence and the risk for premature diabetes and cardiovascular mortality in adulthood. Cardiovascular Diabetology 2018; 17(1): 154.

4. Sörberg Wallin A, Allebeck P, Gustafsson JE, Hemmingsson T. Childhood IQ and mortality during 53 years’ follow-up of Swedish men and women. J Epidemiol Community Health 2018; 72(10): 926–932.

5. Calvin CM, Deary IJ, Fenton C, Roberts BA, Der G, Leckenby N et al. Intelligence in youth and all-cause-mortality: systematic review with meta-analysis. International Journal of Epidemiology 2011; 40(3): 626–644.

6. Hill WD, Marioni RE, Maghzian O, Ritchie SJ, Hagenaars SP, McIntosh AM et al. A combined analysis of genetically correlated traits identifies 187 loci and a role for neurogenesis and myelination in intelligence. Molecular Psychiatry 2019; 24(2): 169–181.

7. Davies G, Lam M, Harris SE, Trampush JW, Luciano M, Hill WD et al. Study of 300,486 individuals identifies 148 independent genetic loci influencing general cognitive function. Nature Communications 2018; 9(1): 2098.

8. Hill WD, Davies G, Liewald DC, McIntosh AM, Deary IJ. Age-Dependent Pleiotropy Between General Cognitive Function and Major Psychiatric Disorders. Biological Psychiatry 2016; 80(4): 266–273.

9. Deary IJ, Weiss A, Batty GD. Intelligence and Personality as Predictors of Illness and Death:How Researchers in Differential Psychology and Chronic Disease Epidemiology Are Collaborating to Understand and Address Health Inequalities. Psychological Science in the Public Interest 2010; 11(2): 53–79.

10. Deary IJ, Harris SE, Hill WD. What genome-wide association studies reveal about the association between intelligence and physical health, illness, and mortality. Current Opinion in Psychology 2019; 27: 6–12.

11. Benyamin B, Pourcain B, Davis OS, Davies G, Hansell NK, Brion MJ et al. Childhood intelligence is heritable, highly polygenic and associated with FNBP1L. Molecular Psychiatry 2013; 19: 253.

12. Pilling LC, Kuo C-L, Sicinski K, Tamosauskaite J, Kuchel GA, Harries LW et al. Human longevity: 25 genetic loci associated in 389,166 UK biobank participants. Aging 2017; 9(12): 2504–2520.

13. Hill W, Marioni R, Maghzian O, Ritchie S, Hagenaars S, McIntosh A et al. A combined analysis of genetically correlated traits identifies 187 loci and a role for neurogenesis and myelination in intelligence. Molecular psychiatry 2018: 1.

14. van Rheenen W, Peyrot WJ, Schork AJ, Lee SH, Wray NR. Genetic correlations of polygenic disease traits: from theory to practice. Nature Reviews Genetics 2019; 20(10): 567–581.

15. Deary IJ. Looking for ‘System Integrity’ in Cognitive Epidemiology. Gerontology 2012; 58(6): 545–553.

16. Davies NM, Hill WD, Anderson EL, Sanderson E, Deary IJ, Davey Smith G. Multivariable two-sample Mendelian randomization estimates of the effects of intelligence and education on health. eLife 2019; 8: e43990.

17. Werme J, van der Sluis S, Posthuma D, de Leeuw CA. LAVA: An integrated framework for local genetic correlation analysis. bioRxiv 2021: 2020.2012.2031.424652.

